# Health economic model to evaluate the cost-effectiveness of smoking cessation services integrated within lung cancer screening

**DOI:** 10.1101/2024.11.27.24318039

**Authors:** Matthew Evison, Rebecca Naylor, Robert Malcolm, Hayden Holmes, Matthew Taylor, Rachael L Murray, Matthew E J Callister, Nicholas S Hopkinson, Sanjay Agrawal, Hazel Cheeseman, David R Baldwin, Zoe Merchant, Patrick Goodley, Alaa Alsaaty, Haval Balata, Philip A.J. Crosbie, Richard Booton

## Abstract

**Introduction:** Integrating smoking cessation support into lung cancer screening can improve abstinence rates. However, healthcare decision makers need evidence of cost effectiveness to understand the cost/benefit of adopting this approach.

**Methods:** To evaluate the cost-effectiveness of different smoking cessation interventions, and service delivery, we used a Markov model, adapted from previous National Institute for Health and Care Excellence guidelines on smoking cessation. This uses long-term epidemiological data to capture the prevalence of the smoking-related illnesses, where prevalence is estimated based on age, sex, and smoking status. Probabilistic sensitivity analysis was conducted to capture joint parameter uncertainty.

**Results:** All smoking cessation interventions appeared cost-effective at a threshold of £20,000 per quality-adjusted life year, compared to no intervention or behavioural support alone. Offering immediate smoking cessation as part of lung cancer screening appointments, compared with usual care (onward referral to stop smoking services) was also estimated to be cost-effective with a net monetary benefit of £2,198 per person, and a saving of between £34 and £79 per person in reduced workplace absenteeism among working age attendees. Estimated healthcare cost savings were more than four times greater in the most deprived quintile compared to the least deprived, alongside a fivefold increase in QALYs accrued.

**Conclusions:** Smoking cessation interventions within lung cancer screening are cost-effective and should be integrated so that treatment is initiated during screening visits. This is likely to reduce overall costs to the health service, and wider integrated care systems, improve quality and length of life, and may lessen health inequalities.

**Key messages:** *What is already known on this topic?:* Smoking cessation interventions are known to be cost-effective in general. However, their cost-effectiveness specifically within lung cancer screening programmes, where they are not routinely commissioned, remains to be established.

*What this study adds:* This health economic analysis estimates that offering smoking cessation immediately within a lung cancer screening visits is a cost-effective intervention, with a substantial return on investment for the healthcare service, alongside a reduction in health inequalities and an increase in productivity for the wider economy.

*How this study might affect research, practice or policy:* This economic evaluation will provide those commissioning and planning healthcare services with evidence that supports the case for funding smoking cessation services integrated within lung cancer screening programmes as immediate, opt-out services.

## INTRODUCTION

Lung cancer is the most common cause of cancer-related deaths worldwide (1). Lung cancer screening has been shown to reduce mortality from both lung cancer and all causes (2). The English Lung Cancer Screening (LCS) programme invites people aged between 55 and 74 who either smoke or used to smoke for a “lung health check” where those assessed as at high risk of lung cancer are offered low-dose computed tomography (LDCT) imaging (3). Attending LCS appointments represents a “teachable moment”(4) to deliver smoking cessation support to participants that actively smoke. Helping smokers to quit has the potential to increase the impact of LCS both by reducing the risk of future cancers and by reducing the risk from other medical conditions such as COPD and cardiovascular disease. Smoking-related disease is also a major driver of health inequalities (5, 6) and contributes to workplace absence due to ill health and early retirement. Approximately one-third of people attending screening programmes in England are current smokers, but dedicated funding for smoking cessation services integrated within the LCS programme is not provided and current national service specifications suggests brief advice is provided during the screening visit with an onward referral to a local stop smoking service. Whilst enhanced smoking cessation interventions are encouraged, no funding stream is available to deliver this. Clinical trial and other data suggest that initiating smoking cessation interventions immediately during screening visits with an ongoing support package afterwards through a single stop smoking service embedded within the screening programme (no onward referral needed) is more effective than onward referral (7–11), increasing the number of people engaged with specialist support and receiving tobacco dependency pharmacotherapy and other interventions. The objective of this study was to develop a health economic model to evaluate the cost-effectiveness of smoking cessation interventions within lung cancer screening participants and immediate initiation (and ongoing delivery) of treatment & support within LCS visits by a single, integrated stop smoking service, to guide healthcare decision makers as to whether this approach would represent good value for money to the NHS.

## METHODS

The economic evaluation was aligned to National Institute for Health and Care Excellence (NICE) guidance (12) using a model structure adapted from a previous economic model on smoking cessation developed as part of its public health guidelines (13). The details of the decision problem are summarised in Table 1. The Markov model structure uses annual cycles where current smokers have a probability of quitting (and becoming ‘former smokers’) and former smokers have a probability of relapsing. We used long-term epidemiological data to model the prevalence of complications associated with the six long-term smoking-related illnesses: lung cancer (LC), coronary heart disease (CHD), chronic obstructive pulmonary disease (COPD), myocardial infarction (MI), stroke, and asthma exacerbation (Figure 1). The model estimates the costs and utilities for each comorbidity using a prevalence-based approach (based on age, gender and smoking status). Throughout the model each health state has an associated utility value, and each comorbidity has an associated cost and disutility associated with the disease occurring. These costs and utilities were applied during each annual cycle and summed to estimate lifetime costs and QALYs across all cycles. The model captures the average lifetime costs, lifetime QALYs, and subsequent cost-effectiveness across all adult populations aged between 55 and 74, consistent with the population eligible for LCS. Average outcomes across the population were calculated by obtaining results for each specific age and applying a weighted average based on the number of people of that age in the population, obtained from 2022 Office for National Statistics (ONS) UK population estimates (14). Costs were applied from the perspective of the England and Wales healthcare system. Outcomes were quantified in terms of quality-adjusted life years (QALYs). Costs and QALYs were discounted at 3.5% per annum. A threshold of £20,000 per QALY gained was used to define cost-effectiveness. The population of the model was defined by the lung cancer screening programme age range (aged 55 to 74). Seven smoking cessation interventions were included within the model (Table 1). Healthcare, societal, and equity outcomes are included within the model as well as lost productivity due to absence from work. To capture the impact on health inequalities, we used the uptake of the lung cancer screening programme by index of deprivation (IMD) to estimate the potential incremental costs and QALYs by IMD group. Smoking can have a lifetime impact, due to the increased risk of long-term health conditions, leading to differences in mortality. As such, a lifetime horizon was used with annual cycles.

**Figure 1:**
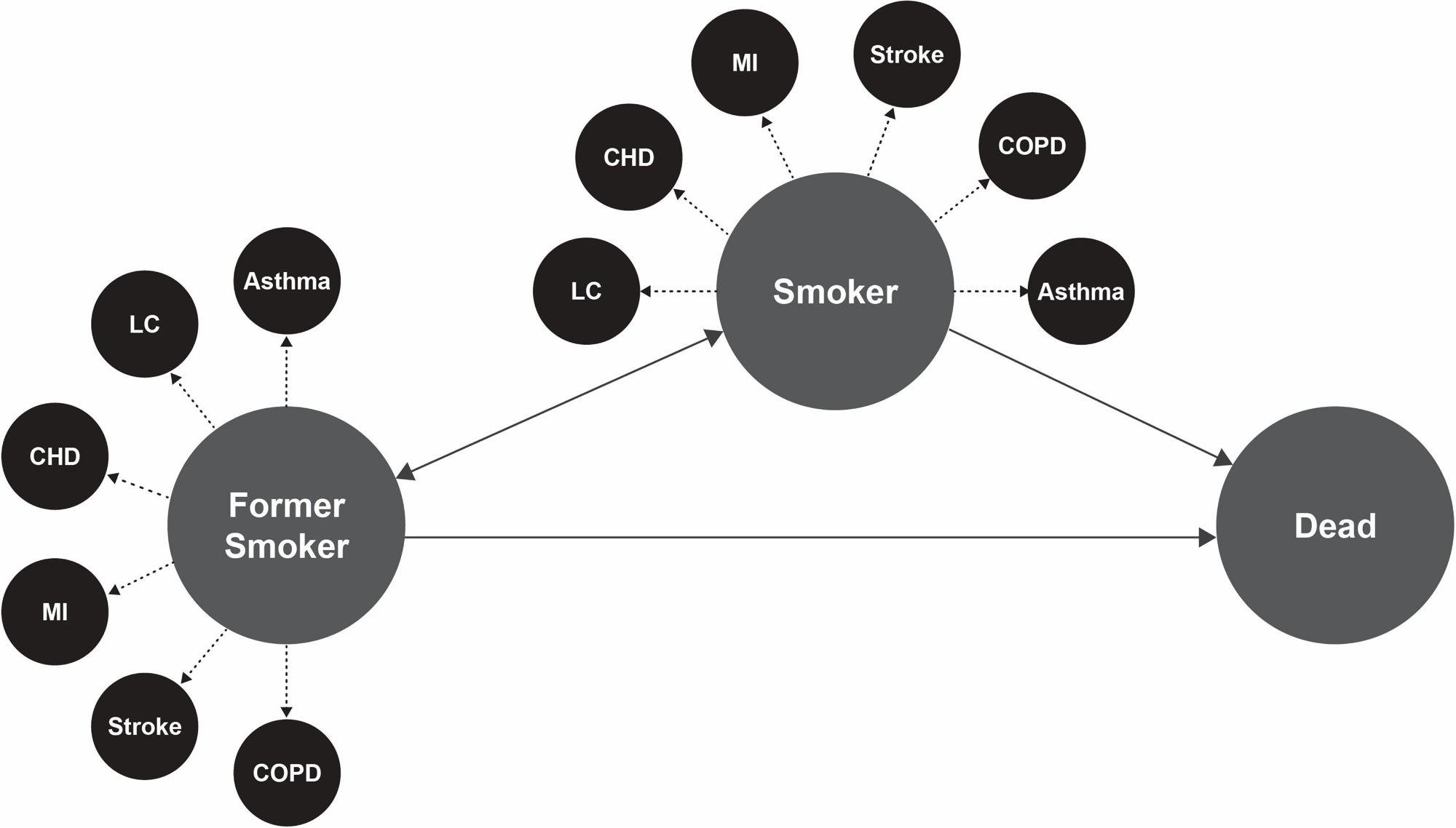
Health economic model structure

**Table 1:** Decision problem.

| Model element | Description |
| --- | --- |
| Population | Adults aged 55 to 74 who currently smoke (eligible for lung cancer screening). |
| Intervention | Smoking cessation interventions:<br>NRT, either long- or short-acting forms ('NRT l/s')<br>NRT, using a combination of long- and short-acting forms ('NRT l&s')<br>Cytisine<br>Varenicline<br>E-cigarettes<br>Varenicline + NRT l/s<br>E-cigarettes + NRT l/s<br>Smoking cessation delivered alongside targeted lung cancer screening (a combination of smoking cessation interventions) |
| Comparator | No intervention or behavioural support only<br>Usual care (referral to stop smoking services) |
| Key outcomes of the model | Economic: Total costs, total QALYs, ICER and net monetary benefit<br>Societal: Lost productivity costs<br>Inequalities: Impact by IMD group (costs and QALYs). |
| Time horizon (years) | Lifetime |

Abbreviations: ICER – Incremental cost-effectiveness ratio, IMD - Index of Multiple Deprivation, NMB – Net monetary benefit, NRT – Nicotine replacement therapy, QALY – Quality adjusted life year.

### Model input parameters

Data used to populate the model were taken from multiple sources. Where required, targeted searches were carried out to identify new data to update parameter values in the existing NG209 model (13). Where parameters remained the same as in the original model, this was because a more appropriate value was not identified from literature searches. The age distribution of the target population was taken from ONS data (15), and aligned with previous studies of people reporting to TLHCs (16). ONS data was used as these were similar to those in lung cancer screening but were stratified by year rather than 5-year age bands. Population gender ratios were also taken from ONS data. The modelling approach also accounted for the likely IMD group weighting of the cohort, taken from Murray et al. (17).

We also considered the impact of usual care, delivered through referral to local stop smoking services, versus smoking cessation delivered immediately and continued through LCS programmes. Uptake of smoking cessation in usual care was provided by Wu et al. (18) and uptake being offered at lung screening by Murray et al. (17). The weighted proportions were provided by Williams et al. (9) in usual care and Murray et al. (17) at lung cancer screening (Supplementary materials Table E1).

### Effectiveness

Effectiveness of interventions was measured by calculating the absolute probability of quitting smoking within six months and probability of quitting within twelve months (Supplementary materials, Table E2).

### HRQoL

Initial health-related quality of life (HRQOL) utilities for smokers, and former smokers were taken from Vogl et al 2012 (19). HRQOL values for smokers and former smokers were also calculated by age. Population age-related utility values were taken from the Health Survey England (HSE) 2014 dataset reported by the NICE Decision Support Unit (20). Utilities were sourced for the included comorbidities, as well as for being a former smoker or smoker, regardless of co-morbidities (Supplementary materials, Table E3).

### Costs

For each intervention, costs were separated into private and healthcare costs, identified using the percentage purchased via prescription (healthcare) and the percentage purchased over the counter (private). Various NRT products were weighted by use to generate the average cost of single and combination therapies. Single mode therapies were weighted according to the percentage quantity (weight of active ingredient i.e. nicotine) of a prescribed product out of all prescribed NRT products, using NHS Prescription Cost Analysis data for 2023/24 (21). Weights for products (from here labelled the product weights) used for combination therapy were calculated from NICE NG209 (13) which used a network meta-analysis (NMA) from Thomas et al 2020 (Supplementary material, Table E4). The total cost of each NRT product was calculated using resource use, unit costs for each dose, and weighted use of each dose (Supplementary materials, Table E5). Annual costs for the included comorbidities were taken from a variety of sources and inflated to 2022/23 prices using the PSSRU inflation index (22) (Supplementary materials, Table E6). A scenario capturing the potential additional costs of implementing the smoking cessation services within the lung cancer screening programme was included in the analysis. This cost was taken from the Yorkshire Enhanced Stop Smoking study [unpublished] which calculated staff cost (on-site stop smoking practitioners present during the screening visit) at £42.60 per participant that currently smokes in 2019/2020 prices. Inflated to 2022/2023 prices using the PSSRU 2024 indices (33) this value is £47.93. This value is included as a scenario rather than in the base case due to opportunity cost of the smoking cessation services. In usual care, if someone was referred for smoking cessation services, they would have an appointment and see clinical staff members. Whether the smoking cessation is initiated in usual care or at lung cancer screening programme, this is likely a similar staff cost, or a redeployment of existing staff time. However, in the case that additional costs are also incurred, this scenario has been run in the analysis. We assumed zero delivery cost attached to receiving no intervention.

### Co-morbidity epidemiology

Epidemiology inputs were required for the included comorbidities. To calculate a gender-weighted risk of each comorbidity by age, prevalence of smokers, former smokers, and non-smokers was also required by gender and age group. These were all taken from the HSE 2022 (23). Non-smokers made up the remaining population. Data sources and risk ratios for each co-morbidity are provided in Supplementary materials, Tables E7-E13)

### Mortality

Mortality inputs were required by age and by smoking status. Mortality by smoking status was split into ten-year age intervals, reported as deaths per 1000 men by Doll et al 1994 (24). These were then converted into risk ratios, for non-smoker and former smokers compared with smokers, where smokers risk ratios were then equal to 1 (Supplementary materials, Tables E14 & E15). Although that study was only focused on a male population, similar evidence has been demonstrated in female populations. Alternative risk ratios were also sourced from the Tobacco Advisory Group of the Royal College of Physicians (25). These were stratified by gender but not age. Mortality by age was taken from ONS life tables for ages 55 to 100 (14).

### Productivity

Productivity was measured by costing absenteeism, calculated by multiplying the excess number of days absent due to smoking by the average daily wage. This required the excess number of days absent, calculating the difference between average annual sick days for non-smokers and average annual sick days for smokers. Average daily wages were calculated for ages 50 and above using average weekly wages for men and women. Average retirement age and proportion of smokers in employment were also required. These data were all taken from the ONS (Supplementary materials, Table E16)

### Sensitivity analysis

Probabilistic sensitivity analysis (PSA) was used in this economic model. The incremental costs and impact on QALYs across all eligible population ages were calculated as a weighted mean based on population averages from the ONS. The PSA results included both pairwise comparisons of different smoking cessation interventions, as well as comparing usual care with smoking cessation provided at the targeted lung cancer screening. The weightings for different interventions (including no intervention) included in usual care and smoking cessation provided at the screening were also varied within PSA (Supplementary materials, Table E17).

## RESULTS

### Treatment-specific smoking cessation analysis

All smoking cessation interventions modelled would lead to substantially more participants abstinent from tobacco at 12-months, bringing reductions in the prevalence of smoking-related diseases and mortality compared with behavioural support alone. All treatments were estimated to be cost-effective compared with behavioural support only, well below the cost-effectiveness threshold of £20,000 per QALY, and all dominated behavioural support alone (Table 2). The biggest impacts of smoking on individuals’ long-term condition costs were related to stroke and CHD. All smoking cessation interventions demonstrated wider benefits to society beyond healthcare costs, improving productivity by reducing absenteeism (Supplementary materials, Table E18). Using a scenario where the time horizon was reduced from lifetime to five years, all the interventions remained cost-effective and dominant compared with behavioural support only, being both more effective and saving the NHS money (Supplementary material, Table E19). Running the model for only those of working age population (aged between 55-65) demonstrated lost productivity costs, with smoking cessation programmes saving between £34 and £79 per person in reduced workplace absenteeism. The PSA showed all interventions are cost-effective in the pairwise comparison (Supplementary materials, Appendix A), with each intervention being cost-effective in above 99% of the PSA iterations compared with behaviour support only (Supplementary materials, Table E20). The PSA results for the fully incremental analysis indicated that the combination of e-cigarettes and NRT is cost-effective in the most iterations (47.6%).

**Table 2:**
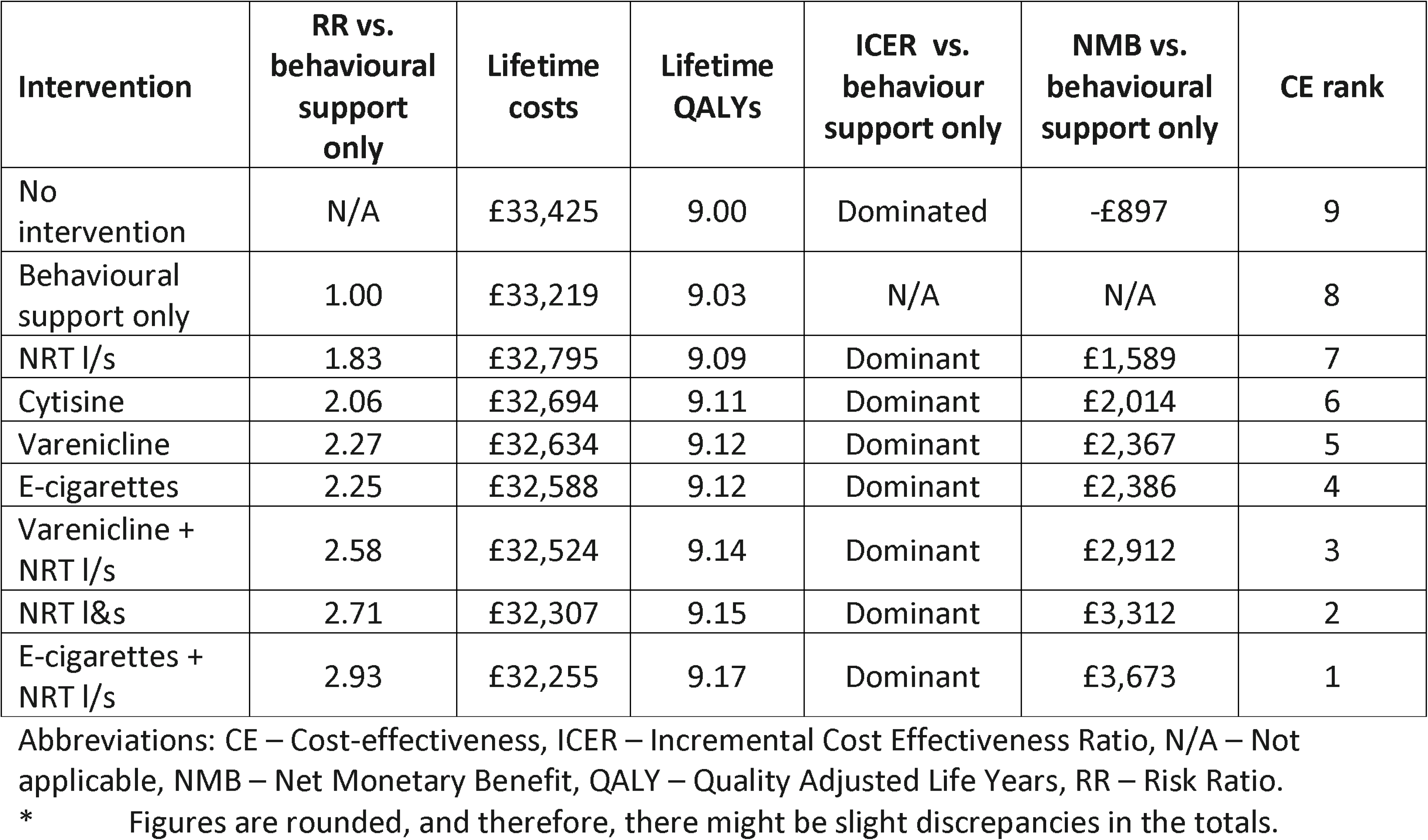
Incremental cost-effectiveness results (per person): fully incremental analysis (lifetime)

### Smoking cessation initiated at lung cancer screening versus usual care of onward referral

The base case analysis suggests that smoking cessation initiated immediately during a lung cancer screening visit and continued through a single stop smoking service integrated within the lung screening service is more effective and less costly than usual care. Regardless of the cost-effectiveness threshold used, the ICER is dominant, with a positive net monetary benefit of £2198 per person (Table 3). This was driven by increased uptake of smoking cessation interventions when initiated immediately during a screening visit, including increased use of pharmacotherapy to support smoking cessation, compared with usual care (onward referral to stop smoking services). A further analysis, where the treatments offered through usual care and the LCS were the same but uptake rates differed, also found smoking cessation initiated through LCS to be more effective and less costly than usual care. Regardless of the cost-effectiveness threshold used, the ICER were dominant, with a positive net monetary benefit of £1,942 per person (Supplementary materials, Table E21). Using a cost-effectiveness threshold of £20,000 per QALY, smoking cessation initiated at targeted lung screening programme was cost-effective in 100% of the 1,000 simulations (Table 4, Figure 2). A scenario was run with the use of e-cigarettes removed from the treatment programme. The ICER is also dominant here with a net monetary benefit of £1679 per person (Table 5). A further scenario was run, where the additional staff cost of providing smoking cessation services at the lung cancer screening programme was included. This is likely an overestimation of the staff cost, and therefore a conservative estimate of the net monetary benefit, given the potential chance of double counting and opportunity cost. The ICER is dominant here, with little change to the NMB at £2150 per person (Table 6). Considering health inequality, for 1,000 smokers attending a national LCS programme, the potential cost savings and QALY gains from integrated smoking cessation interventions for each IMD quintile are presented in Figure 3. Over 4 times the healthcare cost savings would be made (Figure 3A) and over 5 times more QALYs would be accrued (Figure 3B) in the most deprived quintile of the population compared to the least deprived. Finally, a scenario was conducted to determine the time horizon at which point smoking cessation initiated immediately at the lung cancer screening visits became cost-effective. The results suggested that a time horizon of 3 years or greater estimates a positive net monetary benefit, and a cost-effective intervention (Supplementary material, table E22).

**Figure 2:**
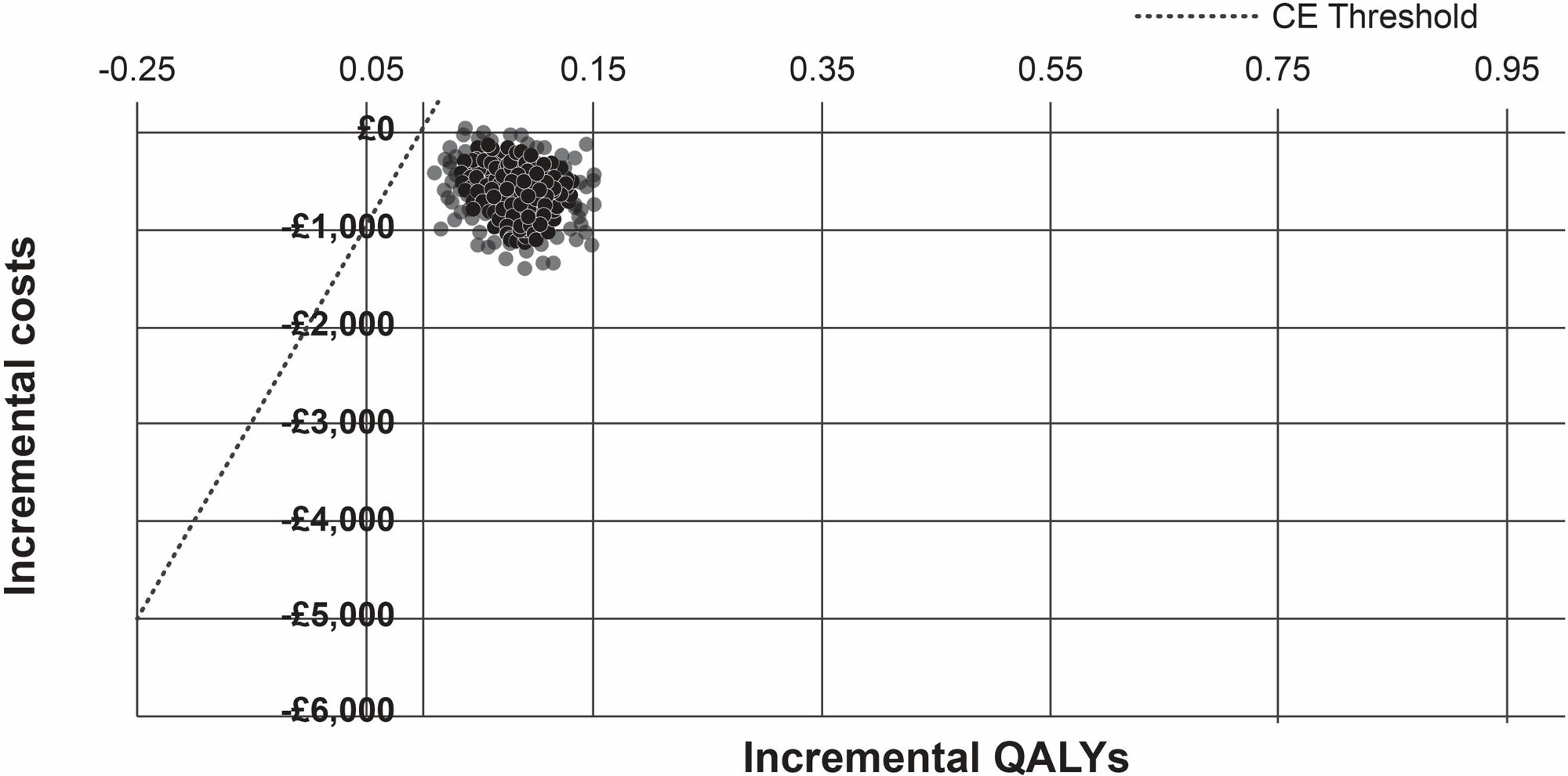
Cost-effectiveness plane: smoking cessation initiated at lung cancer screening versus usual care of onward referral

**Figure 3A:**
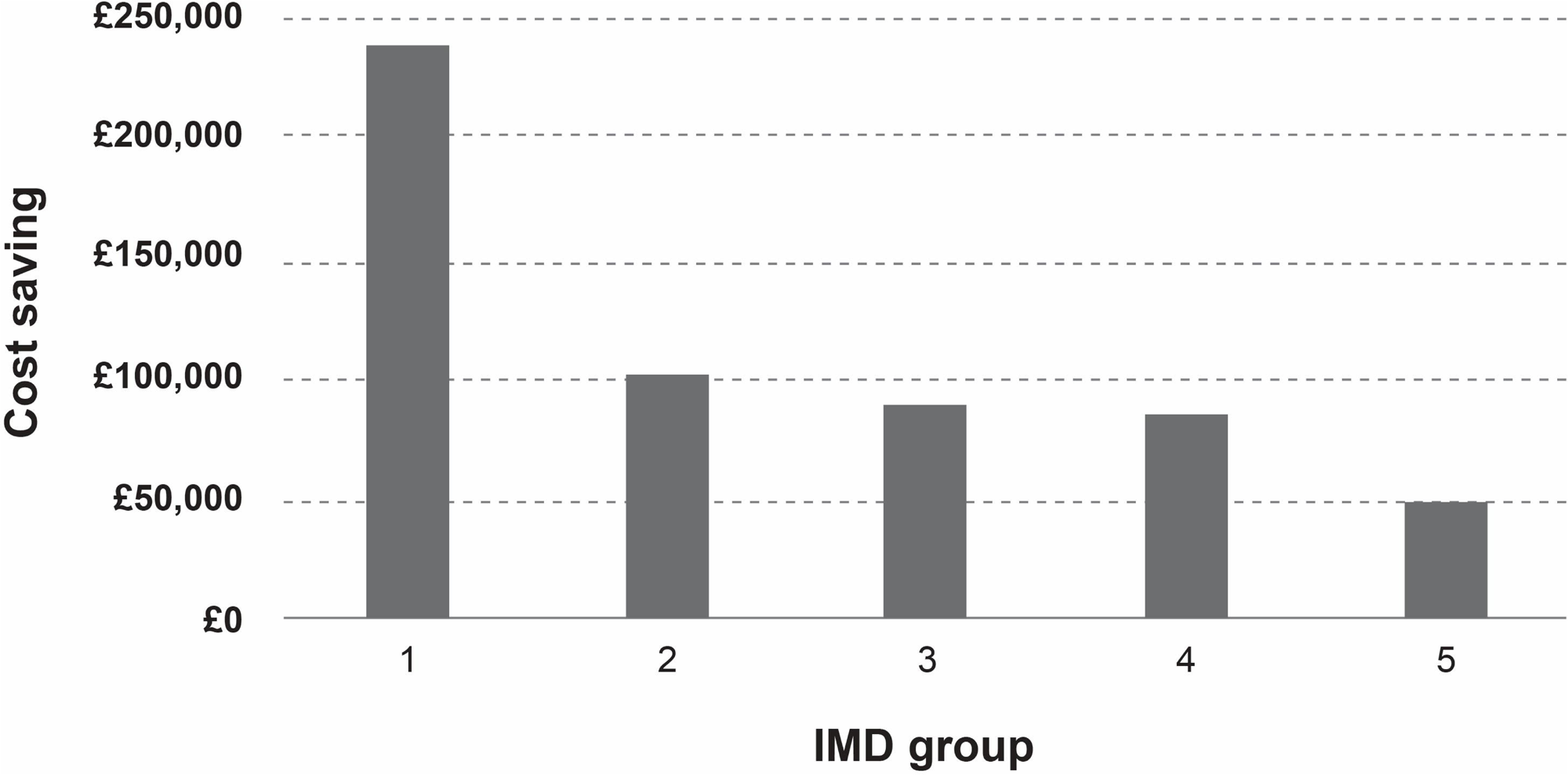
Incremental cost saving per IMD quintile per 1000 people that smoke attending lung cancer screening.

**Figure 3B:**
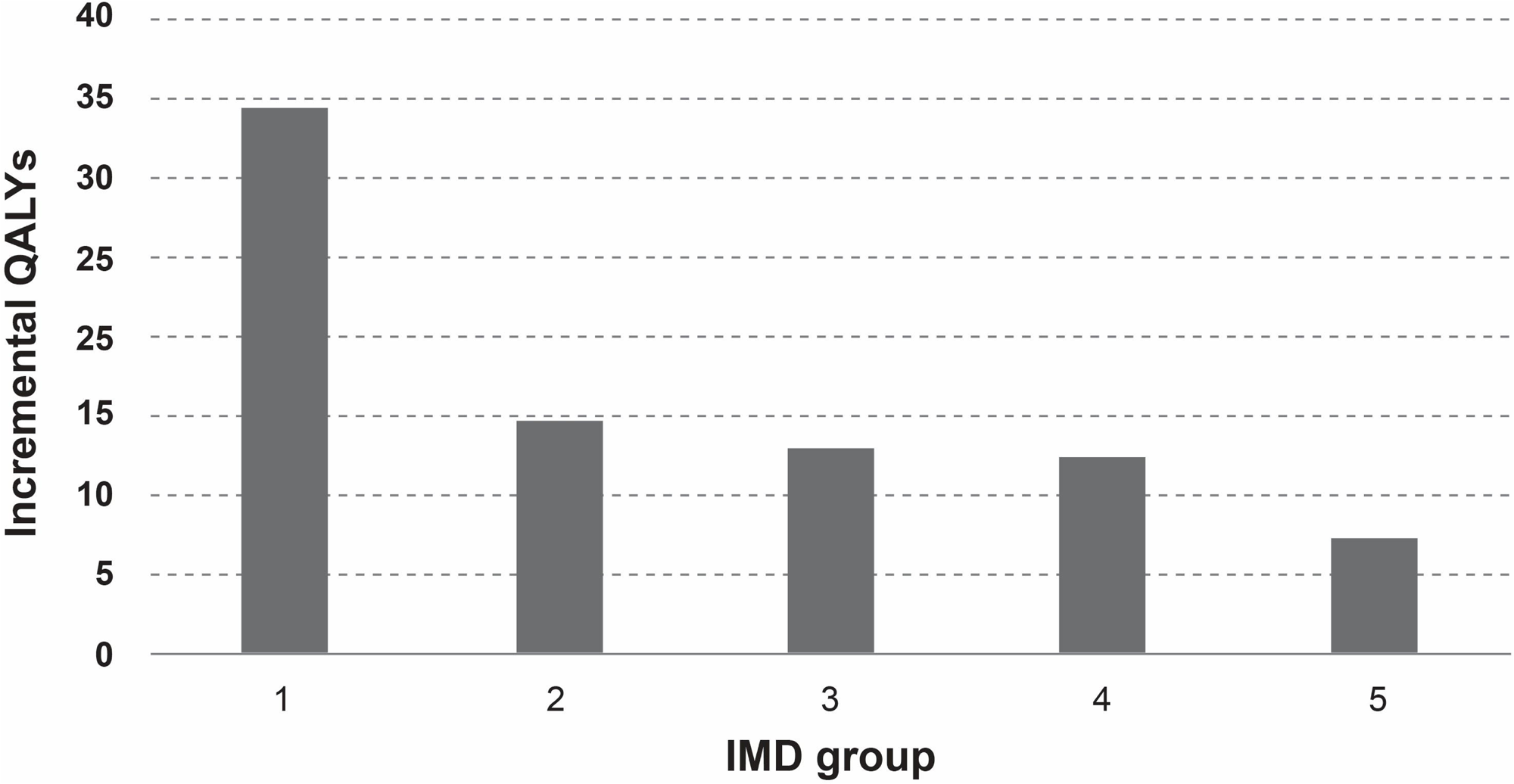
Incremental Qalys per IMD quintile per 1000 people that smoke attending lung cancer screening

**Table.** Error! No text of specified style in document.: Summary of deterministic base case results for smoking cessation initiated at lung cancer screening versus usual care of onward referral.

| Result | Smoking cessation initiated at targeted lung screening programme | Usual care | Incremental |
| --- | --- | --- | --- |
| Costs per person | £32,779 | £33,344 | -£564 |
| QALYs per person | 9.09 | 9.01 | 0.08 |
| Private costs | £67 | £1 | £67 |
| Lost productivity costs | £284 | £314 | -£30 |
| ICER |  |  | Dominant |
| Net monetary benefit |  |  | £2,198 |

\* Figures are rounded, and therefore, there might be slight discrepancies in the totals.

**Table 4:** Probabilistic sensitivity results for smoking cessation initiated at lung cancer screening versus usual care of onward referral.

| Result | Smoking cessation initiated at targeted | Usual care | Incremental |
| --- | --- | --- | --- |

|  | lung screening programme |  |  |
| --- | --- | --- | --- |
| Total cost | £33,156 | £33,769 | -£613 |
| Total QALYs | 9.01 | 8.92 | 0.09 |
| Incremental cost-effectiveness ratio |  |  | **Dominant** |
| Net monetary benefit |  |  | **£2,326** |
| Probability of cost-effective (£20,000 per QALY) |  |  | **100%** |

\* Figures are rounded, and therefore, there might be slight discrepancies in the totals.

**Table 5:** Summary of deterministic base case results for smoking cessation initiated at lung cancer screening versus usual care of onward referral, where e-cigarettes are removed from the treatment programme.

| Result | Smoking cessation initiated at targeted lung screening programme | Usual care | Incremental |
| --- | --- | --- | --- |
| Costs per person | £32,940 | £33,371 | -£431 |
| QALYs per person | 9.07 | 9.01 | 0.06 |
| Private costs | £24 | £1 | £24 |
| Lost productivity costs | £293 | £315 | -£23 |
| ICER |  |  | **Dominant** |
| Net monetary benefit |  |  | **£1,679** |

**Table 6:** Summary of deterministic base case results for smoking cessation initiated at lung cancer screening versus usual care of onward referral, with additional staff cost for delivering service.

| Result | Smoking cessation initiated at targeted lung screening programme | Usual care | Incremental |
| --- | --- | --- | --- |
| Costs per person | £32,827 | £33,344 | -£516 |
| QALYs per person | 9.09 | 9.01 | 0.08 |
| Private costs | £67 | £1 | £67 |
| Lost productivity costs | £284 | £314 | -£30 |
| ICER |  |  | **Dominant** |
| Net monetary benefit |  |  | **£2,150** |

## DISCUSSION

The key finding of this analysis is that smoking cessation treatment integrated within LCS is cost-effective. All smoking cessation interventions evaluated were estimated to be cost effective as all resulted in healthcare savings and additional QALYs, dominating behavioural support only or no intervention. Smoking cessation treatment and support, initiated immediately at the point of lung cancer screening and continued by a single stop smoking team integrated within the screening programme, would deliver an additional 80 QALYs per 1,000 smokers, and a net monetary benefit to the NHS of £2,198 per person, compared to usual care of onward referral to a stop smoking service. This service delivery model becomes cost-effective with a three-year time horizon. Smoking cessation within lung cancer screening would also increase economic productivity by reducing absence from work. Finally, healthcare cost savings would be over 4 times greater in the most deprived quintile when compared to the least deprived, alongside a corresponding 5 times increase in QALYS accrued, reducing health inequality.

### Significance of findings

The cost-effectiveness of smoking cessation interventions is driven by their effectiveness (the relative quit rates); higher numbers of non-smokers at 12-months when compared to behavioural support only. Increasing the number of quitters at 12-months reduced the number of smokers throughout the remainder of the economic model, reducing the health and economic burdens associated with smoking, through reductions in smoking-related diseases and mortality. All methods of smoking cessation are cost-effective options for smokers attending a lung cancer screening programme. Pharmacotherapies are estimated to deliver the highest health gains and reduce the most costs. Current evidence indicates that pharmacotherapy may be used less in stop smoking services, with a greater uptake of behavioural support only. Combining behavioural support with pharmacotherapy treatments is up to three times more likely to be effective (26). In contrast to costs of smoking-related diseases, intervention costs had a relatively minor influence on the cost-effectiveness results. The total intervention costs across the population are modest when compared with the lifetime net monetary benefit associated with those who quit smoking due to the cessation interventions. For example, intervention costs for cytisine are £115 per person more than behavioural support only. The net monetary benefit for cytisine compared with behavioural support only is £2,014. This would mean that the costs for cytisine could be over 17 times higher and the intervention would still be considered cost-effective compared with behaviour support only.

Previous analysis has indicated superior quit rates when providing smoking cessation immediately during LCS and continued afterwards by a single stop smoking service integrated within the screening programme (7–9, 11). Our analysis has not modelled the direct effectiveness of quit rates from these smaller studies but used a larger meta-analysis of smoking treatments. Studies capturing the effectiveness of immediate initiation of smoking cessation highlight a greater use of pharmacotherapy in the intervention arm, as well as more people taking up smoking cessation support programmes within the intervention arm.

Reducing smoking prevalence should reduce health inequalities, given the link between socioeconomic status and smoking prevalence (27). Based on the uptake of smoking cessation and attendance at screening, delivering smoking cessation interventions within LCS are likely to have the greatest impact in the most deprived populations. Previous evidence looking at wider smoking cessation programmes, including younger adults, has demonstrated that inequalities may increase due to higher uptake and benefits of quitting among more advantaged groups (28). Despite this, authors also suggested the benefits to population health would offset impacts to health inequalities, based on estimated aversion parameters. Furthermore, the population of the cohort used in this analysis is different to previous analysis. For example, the people attending screening were more likely to be from deprived areas than the population considered in the previous analysis population. We believe this should be explored further to determine the true impact on health inequalities.

Further economic benefits of smoking cessation interventions include increased productivity, due to reduced absenteeism for those of working age as well as greater ability to perform childcare and other informal caring in retirement. These benefits are not captured directly in the cost-effectiveness estimates, but the improvements to productivity reflect some of the wider societal benefits to effective smoking cessation programmes and optimising their delivery.

Results from this economic modelling report on pairwise treatments compared with no intervention or behavioural support only were comparable to results reported by studies in the existing literature. The BENESCO model is a common Markov model that is used to estimate the lifetime cost and benefits of smoking. The BENESCO model has been applied across a variety of populations including the USA, the Caribbean, central America, and Europe (29–31). Further papers have considered the different ways smoking cessation programmes could be modelled, either using Markov models, or discrete event simulations (32). In summary, the model structures themselves did not influence smoking cessation cost-effectiveness results, but long-term assumptions did. When there is variation in long-term predictions between interventions, economic models need a structure that can reflect this. Overall, the direction of the results was similar, indicating that smoking cessation is cost-effective regardless of the modelling approach. The multi-treatment results of this analysis are also consistent with Cao et al (33), who modelled the potential impact of cessation interventions at the point of lung cancer screening. This analysis was from a US perspective, however, the exploratory analysis indicated that uptake rates and cessation effectiveness were the key drivers of the results. We believe that smoking cessation effectiveness will be driven by the interventions offered by clinicians, which is reflected in our analysis, and is highlighted as a key driver. All previous studies indicated that the estimates were likely to be more conservative, due to other factors from stopping smoking which were omitted from the analysis. Therefore, the true effect is likely to be greater than the economic model estimates.

### Limitations

The complex impact of smoking and data availability necessitated some assumptions surrounding the clinical pathway and how events are captured (Supplementary materials, Table E22). This model is adapted from NICE guidelines, and key assumptions and their validity were discussed with clinical experts during guideline development. In the base case analysis, e-cigarettes were found to be cost-effective and dominant compared with behavioural support only. However, there is more data needed on the long-term safety of e-cigarettes. The analysis indicates that even if e-cigarettes only reduce the risk of mortality and other chronic conditions by half, when compared with smoking, it would still be a cost-effective intervention. Furthermore, removing e-cigarettes from an integrated smoking cessation programme within LCS reduces the net monetary benefit from £2198 to £1679 per person.

The exclusion of some factors from our model means that the current analysis is likely to be underestimating the real benefits of providing smoking cessation support in this context. The health effects of smoking cessation extend far beyond the six illnesses included, including a higher risk of dementia, other types of cancer, respiratory infections, chronic kidney disease, osteoporotic fractures, mental health conditions, and other high-consequence diseases (25), as well as improved recovery, fewer complications, and shorter hospital stays following surgery (34)(35). A further benefit, not quantified in this analysis is the wider societal impact that smoking cessation can have. Quitting smoking does not just help the smoker themselves, but also those around them through a reduction in second-hand smoke exposure, which can be especially harmful to children (36). Smoking cessation may have a positive impact on other people’s smoking behaviour (37–39). Finally, the time horizon within which the integrated and immediate smoking cessation service becomes cost-effective (three years) is also highly likely to be an under-estimation given that the focus of the model is on long-term conditions and mortality and other healthcare resource use may have been omitted, such as GP appointments or hospitalisations in the short-term.

### Conclusion

This economic analysis suggests that smoking cessation interventions for LCS participants are highly cost-effective. Furthermore, integrating smoking cessation services within the screening programme to initiate treatment at screening visits and continue support afterwards is superior to onward referral to separate stop smoking services.

## Data Availability

All data produced in the present work are contained in the manuscript

## DATA SHARING

There are no data sharing elements to this work.

## AUTHOR CONTRIBUTION

ME and RB conceptualised this study. RN, RM, HH & MT adapted the existing smoking cessation model used in previous NICE guidelines for the lung cancer screening population. RN, RM, HH and MT completed the health economic analysis. ME, RM, BD, MC, SA, HC and NH formed a clinical reference group to review the findings of the evaluation and make adaptations to the model as required. ME, RN, RM and HH led the writing of a first manuscript draft and all authors provided expert review and amendments over several revisions to the final version.

## TRANSPARENCY DECLARATION

Matthew Evison, the manuscript’s guarantor affirms that the manuscript is an honest, accurate, and transparent account of the study being reported; that no important aspects of the study have been omitted; and that any discrepancies from the study as planned (and, if relevant, registered) have been explained.

