## Supplementary materials for "Health economic model to evaluate the cost-effectiveness of smoking cessation services integrated within lung cancer screening"

Supplementary materials, Table E1: Input parameters on population characteristics, uptake of smoking cessation and multi-treatment weightings

| Input | Value | Source |
| --- | --- | --- |
| **IMD weighting** | | |
| IMD1 | 42.0% | Murray et al. (1)  Table 1 |
| IMD2 | 18.0% |  |
| IMD3 | 15.8% |  |
| IMD4 | 15.3% |  |
| IMD5 | 8.9% |  |
| **Uptake of smoking cessation** | | |
| Usual care | 17.4% | Wu et al. (2)  Table 3 |
| Offered at lung screening | 74.8% | Murray et al. (1) |
| **Multi-treatment weightings: Usual care** | | |
| Behavioural support only | 57.5% | Williams et al. (3)  Table 2 |
| NRT l/s | 5.5% |  |
| NRT l&s | 3.8% |  |
| Cytisine | 0.0% |  |
| Varenicline | 2.7% |  |
| E-cigarettes | 30.5% |  |
| Varenicline + NRT l/s | 0.0% |  |
| E-cigarettes + NRT l/s | 0.0% |  |
| **Multi-treatment weightings: Initiated at targeted lung screening programme** | | |
| Behavioural support only | 14.6% | Murray et al. (1)  24% opted for NRT alone (assumed 50:50 split between NRT l/s and NRT l&s  33% combination of NRT and e-cigarette  Pharmacotherapy selected for 7% so this is spread evenly between varenicline and cytisine (3.5%)  4% of respondents had missing data, so all proportions are scaled up to only include available data |
| NRT l/s | 12.5% |  |
| NRT l&s | 12.5% |  |
| Cytisine | 3.6% |  |
| Varenicline | 3.6% |  |
| E-cigarettes | 18.8% |  |
| Varenicline + NRT l/s | 0.0% |  |
| E-cigarettes + NRT l/s | 34.4% |  |

Abbreviations: IMD - Index of multiple deprivation, NRT – Nicotine replacement therapy.

Supplementary materials, Table E2: Input parameters on smoking cessation effectiveness

| Input | Value | Source |
| --- | --- | --- |
| RR of abstinence at 6 months versus usual care for NRT single mode | 1.83 | NICE NG209 Evidence Review K (4)  Table 7 |
| RR of abstinence at 6 months versus usual care for NRT multi-mode | 2.71 |  |
| RR of abstinence at 6 months versus usual care for cystine | 2.06 | Lindson et al 2023 (5)  Converted OR to RR using formula: RR = OR/(1 – p(p*OR)  OR = 2.21; p = 0.06. |
| RR of abstinence at 6 months versus usual care for varenicline | 2.27 | NICE NG209 Evidence Review K (4)  Table 7 |
| RR of abstinence at 6 months versus usual care for e-cigarettes | 2.25 |  |
| RR of abstinence at 6 months versus usual care for varenicline plus NRT single mode | 2.58 |  |
| RR of abstinence at 6 months versus usual care for e-cigarettes plus NRT single mode | 2.93 |  |
| Probability of quitting at six months when referred to stop smoking services (usual care) | 11.5% | Thomas et al 2020  Calculated as r/n across placebo arms in included RCTs. = 3,232/28,139.  Summed total for Figure 1, 6, 9, 13, 18, and 21 in NICE NG209 Evidence Review K. |
| Relapse rate between six months and twelve months | 14.4% | Coleman et al 2010 (6)  Graph 2.1 timepoints: 30.6% quit at 6 months; 26.2% quit at 12 months.  Remaining quitters = 26.2/30.6 = 85.6%. Relapse rate = 100 – 85.6. |
| Natural quit rate (annual background rate) | 5.0% | Livingstone-Banks et al 2019 (7)  Control quit rate average |

Abbreviations: NRT: nicotine replacement therapy; OR: odds ratio; RCT: randomised controlled trial; RR: risk ratio.

Supplementary materials, Table E3: Input parameters for utilities

| Input | Value | Source |
| --- | --- | --- |
| HRQOL of a non-smoker (according to Vogl et al) | 0.88 | Vogl et al 2012 |
| HRQOL of a smoker | 0.85 | Vogl et al 2012  Non-smoker utility minus the average disutility of low, medium, and high-level smokers. |
| HRQOL of a former smoker | 0.87 | Vogl et al 2012 |
| HRQOL value following a stroke | 0.48 | Tengs and Wallace 2000 (8)  Average stroke health states |
| HRQOL value with lung cancer | 0.61 | Bolin et al 2006 (9) |
| HRQOL value following myocardial infarction | 0.80 | Tengs and Wallace 2000  Average of 85 MI health states |
| HRQOL value with coronary heart disease | 0.76 | Stevanović et al 2016 (10)  Value of EQ-5D studies (22) for CHD dataset |
| HRQOL value with COPD | 0.73 | Rutten-van Molken et al 2006 (11) |
| HRQOL value with asthma | 0.73 | Szende et al 2004 (12)  Mapped EQ-5D score from AQLQ score. Exacerbations can last from minutes to days. Assumed utility applies for 1 week. |

Abbreviations: CHD: coronary heart disease; COPD: chronic obstructive pulmonary disease; HRQOL: health-related quality of life; MI: myocardial infarction.

**Supplementary materials, Table E4:** Costs (NRT product weights)

| Input | Value | Source |
| --- | --- | --- |
| Single Mode NRT | | |
| Patch | 24.5% | NHS Prescription Cost Analysis 2023/24 (13)  Weights identified according to the percentage quantity (weight of active ingredient i.e. nicotine) of all prescribed NRT products. |
| Lozenge | 20.1% |  |
| Gum | 27.2% |  |
| Spray | 9.4% |  |
| Inhalator | 18.8% |  |
| Multi-Mode (Combination) NRT | | |
| Patch | 100.0% | Thomas et al 2020 [NICE NG209] (4)  Calculated by r/n where:  r = total number of participants receiving each product as part of a combination NRT strategy for RCTs included in the NMA.  n = total number of participants in the NRT combination arms across all RCTs included in the NMA. |
| Lozenge | 0.0% |  |
| Gum | 50.7% |  |
| Spray | 25.5% |  |
| Inhalator | 5.7% |  |
| Any short acting NRT (lozenge, gum, spray, inhalator) | 18.1% |  |

Abbreviations: NMA: network meta-analysis; NRT: nicotine replacement therapy; RCT: randomised controlled trial.

**Supplementary materials, Table E5:** Costs (Annual product costs)

| Input | Value | Source |
| --- | --- | --- |
| NRT patch | £114.45 | Dosage – BNF 2024:  More than 10 cigarettes per day – high strength patch for 6-8 weeks (7 weeks midpoint), followed by medium strength for 2 weeks, followed by low strength for 2 weeks.  Less than 10 cigarettes per day – medium strength patch for 6-8 weeks (7 weeks midpoint), followed by low strength for 2-4 weeks (3 weeks midpoint).  Unit costs per 7 patches – BNF 2024:  21mg/24hr = £9.97; 14mg/24hr = £10.82; 7mg/24hr = £10.49. Total average dose = £109.81.  25mg/16hr = £11.43; 15mg/16hr = £11.43; 10mg/16hr = £11.43. Total average dose = £120.02.  Weights for 16hr vs 24hr patch usage – Prescription Cost Analysis (2023/24):  24hr patch = 54.55%; 16hr patch = 45.45%. |
| NRT lozenge | £70.31 | Dosage – Schnoll et al 2011 (14):  9 lozenges per day for first six weeks, 5 per day for weeks 7-9, and 3 per day for weeks 10-12.  Total of 546 lozenges.  Unit costs – BNF 2024:  1/1.5mg = £0.12; 2mg = £0.13; 4mg = £0.13.  Weights by strength – Prescription cost analysis (2023/24):  4mg = 29.21%; 2mg = 47.07%; 1/1.5mg = 23.72%. |
| NRT gum | £112.28 | Dosage:  Ad lib when symptoms occur. Assumed intake of 1 per 1.5 hours over 16 waking hours = 12 per day. 12-week treatment duration assumption.  Unit costs – BNF 2019 (no longer listed in BNF):  2mg = £0.09; 4mg = £0.11. Inflated from 2019 to 2022/23 prices using PSSRU (15).  Weights by strength – Prescription cost analysis 2018:  4mg = 45.00%; 2mg = 55.00%. |
| NRT spray | £87.58 | Dosage – NRT Consumer Medicine Information 2013 (16):  Weeks 1-6: 1-2 sprays when normally smoke a cigarette. Weeks 7-9: reduce number of sprays per day, by end of week 9 should be using half used in step 1. Weeks 10-12: continue reducing so no more than 4 sprays per day are used during week 12 [64 sprays per day max.]. Average cigarettes per day in UK is 10.6 (ONS 2024 (17)). Average of 1014.3 sprays over 12 weeks (1.5 sprays per cigarette.  Unit costs – BNF 2024:  Nasal = £21.05 per pack; oral = £19.13 per pack.  Weights by spray type – Prescription cost analysis 2023/24:  Nasal = 6.5%; oromucosal = 93.5%. |
| NRT inhalator | £427.90 | Dosage – Nicorette 2023 (18):  Take as required. Individuals should not exceed 6 cartridges of 15mg strength daily. Recommended that 3-6 15mg cartridges used daily (midpoint 4.5 cartridges daily).  Assumed 12-week duration.  Unit costs – BNF 2024:  Per 15mg cartridge = £1.13 (based on 20 cartridges). |
| Any short acting NRT | £185.19 | Prescription cost analysis 2023/24 (13)  Weighted costs of lozenge, gum, spray and inhalator.  Weights identified according to the percentage quantity (weight of active ingredient i.e. nicotine) of all prescribed short acting NRT products. |
| Cytisine | £115.00 | BNF 2024  Dosage: 1.5mg every 2 hours from day 1-3, maximum 9mg per day. Then reduce to 1.5mg every 2.5 hours from day 4-12. Smoking should be stopped no later than day 5. Reduce to 1.5mg every 3 hours days 13-16, reduce to 1.5mg every 5 hours days 17-20, reduce to 1.5-3mg per daily days 21-25.  Cost per 100 pack of 1.5mg tablets = £115. |
| Varenicline | £181.94 | BNF 2019  Dosage: one 0.5mg tablet for 3 days, 0.5mg twice daily for 4 days, 1mg tablet twice day for 11 weeks. Cost per tablet (0.5mg and 1mg) - £0.98 (multiplied by 165 tablets).  Inflated from 2019/20 to 2022/23 prices using PSSRU indices (15), since no longer listed on BNF. |
| E-cigarette starter pack and replacement coils | £33.98 | Hajek et al 2022 (19), and UK Ecig Store 2024 (20)  Hajek uses on kit by UK E-cig store which included start kit and 5 replacement coils.  2024 price = £24.99 for starter kit, £8.99 for the replacement coils. |
| E-liquid | £223.44 | UK Ecig Store 2024 (20)  Average cost of 10ml fluid = £3.99.  10mp fluid will last 3-10 days (midpoint 6.5). Assume intake isn’t reduced over a year. |

Abbreviations: BNF – British National Formulary, ONS – Office For National Statistics, PSSRU - Personal Social Services Research Unit, UK – United Kingdom.

**Supplementary materials, Table E6:** Costs (Co-morbidity costs)

| Input | Value | Source |
| --- | --- | --- |
| Annual cost of one stroke | £7,123 | Xu et al 2018 (21)  Table 2  Mean total health and social care costs = £46,039 Year 5) - £22,429 (year 1) = £23,610 (four subsequent years).  £23,610/4 = £5,902.50 = annual cost.  Inflated from 2015/16 to 2022/23 prices using PSSRU 2024 indices (15). |
| Annual cost of one person with lung cancer | £11,211 | Cancer Research UK 2012 (22)  Costs UK health systems £9.071. Inflated from 2012/13 to 2022/23 prices using PSSRU 2024 indices (15). |
| Annual cost of one myocardial infarction | £2,995 | Danese et al 2016 (23)  Table 2  First and second cost combined, annualised future costs = £2,472.28.  Inflated from 2014/15 to 2022/23 prices using PSSRU 2024 indices (15). |
| Annual cost of one person with coronary heart disease | £6,355 | Landeiro et al 2024 (24)    Report cost of £5,530 per person.  Inflated from 2018/19 to 2022/23 prices using PSSRU 2024 indices (15). |
| Annual cost of one person with COPD | £1,914 | Punekar et al 2014 (25)  "Total annual per patient COPD costs were £1,523, £2,405, and £3,396 for patients with no, one, and two or more exacerbations, respectively"  Assumed conservatively with no moderate/severe exacerbations (£1,523).  Inflated from 2011/12 to 2022/23 prices using PSSRU 2024 indices (15). |
| Annual cost of one asthma exacerbation | £1,492 | Leaviss et al 2014 (26)  Reported cost of £1,162.25.  Inflated from 2010/11 to 2022/23 prices using PSSRU 2024 indices (15). |

Abbreviations: COPD – Chronic Obstructive Pulmonary Disease, PSSRU - Personal Social Services Research Unit, UK – United Kingdom.

**Supplementary materials, Table E7:** Prevalence of smokers by age and sex

| Age range | Non-smoker | Former smoker | Smoker | Source |
| --- | --- | --- | --- | --- |
| Males | | | | |
| 55 to 64 | 57.6% | 30.0% | 12.4% | HSE 2022 (27) |
| 65 to 74 | 55.3% | 34.6% | 10.1% |  |
| 75+ | 52.6% | 42.6% | 4.8% |  |
| Females | | | | |
| 55 to 64 | 64.0% | 24.2% | 11.8% | HSE 2022 (27) |
| 65 to 74 | 58.9% | 30.9% | 10.1% |  |
| 75+ | 62.9% | 31.9% | 5.2% |  |

Abbreviations: HSE: Health Survey for England.

**Supplementary materials, Table E8:** Stroke prevalence and RR

| Input | Value - males | Value - females | Source |
| --- | --- | --- | --- |
| Prevalence 55 to 64 | 2.7% | 2.0% | Bhatnagar et al 2014 (28)  Table 2 |
| Prevalence 65 to 74 | 6.4% | 4.4% |  |
| Prevalence 75+ | 14.9% | 12.4% |  |
| Risk ratio – smokers to non-smokers | 1.47 | 2.03 | Myint et al 2008 (29)  Table 3 |
| Risk ratio – former smokers to non-smokers | 1.00 | 1.02 |  |

**Supplementary materials, Table E9:** Lung cancer prevalence and RR

| Input | Value - males | Value - females | Source |
| --- | --- | --- | --- |
| Prevalence 55 to 64 | 0.089% | 0.076% | Maddams et al 2008 (30)  Table 4 |
| Prevalence 65 to 74 | 0.748% | 0.331% |  |
| Prevalence 75+ | 0.748% | 0.331% |  |
| Risk ratio – smokers to non-smokers | 8.78 | 7.48 | O’Keeffe et al 2018 (31)  Supplementary Figure 1 |
| Risk ratio – former smokers to non-smokers | 3.01 | 2.82 |  |

**Supplementary materials, Table E10:** Myocardial infarction prevalence and RR

| Input | Value - males | Value - females | Source |
| --- | --- | --- | --- |
| Prevalence 55 to 64 | 4.5% | 0.9% | HSE 2017 (32)  Table 1 |
| Prevalence 65 to 74 | 11.6% | 3.4% |  |
| Prevalence 75+ | 12.9% | 5.1% |  |
| Risk ratio – smokers to non-smokers | 2.23 | 3.46 | Millett et al 2018 (33)  Figure 1 |
| Risk ratio – former smokers to non-smokers | 1.20 | 1.25 |  |

**Supplementary materials, Table E11:** Coronary heart disease prevalence and RR

| Input | Value - males | Value - females | Source |
| --- | --- | --- | --- |
| Prevalence 55 to 64 | 6.1% | 2.3% | British Heart Foundation 2024 (34)  Chapter 2, Sheet 2.19  Averages were calculated for the two five-year age groups that fell into each ten-year age group. |
| Prevalence 65 to 74 | 12.9% | 5.4% |  |
| Prevalence 75+ | 25.1% | 14.7% |  |
| Risk ratio – smokers to non-smokers | 1.60 | 1.70 | Shields et al 2013 (35) |
| Risk ratio – former smokers to non-smokers | 1.10 | 1.40 |  |

**Supplementary materials, Table E12:** COPD and RR

| Input | Value - males | Value - females | Source |
| --- | --- | --- | --- |
| Prevalence 55 to 64 | 4.2% | 4.2% | Public Health England 2011 (36) |
| Prevalence 65 to 74 | 8.3% | 8.3% |  |
| Prevalence 75+ | 8.9% | 8.9% |  |
| Risk ratio – smokers to non-smokers | 4.11 | 3.28 | Forey et al 2011 (37)  Table 7 |
| Risk ratio – former smokers to non-smokers | 2.87 | 2.02 |  |

**Supplementary materials, Table E13:** Asthma prevalence and RR

| Input | Value - males | Value - females | Source |
| --- | --- | --- | --- |
| Smokers | | | |
| Incidence 55 to 64 | 0.05% | 0.05% | Leaviss et al 2014 (reporting from Asthma UK 2004) (26) |
| Incidence 65 to 74 | 0.07% | 0.06% |  |
| Incidence 75+ | 0.07% | 0.06% |  |
| Long-term quitters | | | |
| Incidence 55 to 64 | 0.05% | 0.05% | Leaviss et al 2014 (reporting from Asthma UK 2004) (26) |
| Incidence 65 to 74 | 0.06% | 0.05% |  |
| Incidence 75+ | 0.06% | 0.05% |  |

**Supplementary materials, Table E14:** Mortality per 1000 men

| Age | Non-smoker | Former smoker | Smoker |
| --- | --- | --- | --- |
| 55 to 64 | 9.50 | 13.40 | 20.30 |
| 65 to 74 | 23.70 | 31.60 | 47.00 |
| 75 to 84 | 67.40 | 77.30 | 106.00 |
| 85+ | 168.60 | 179.70 | 218.70 |
| Source: | Doll et al 1994 (38) Table IV | | |

**Supplementary materials, Table E15:** Alternative mortality RRs

|  | Former Smoker | Smoker |
| --- | --- | --- |
| Male | 1.27 | 2.66 |
| Female | 1.06 | 2.23 |
| Source: | Tobacco Advisory Group of the Royal College of Physicians 2018 (39)  Section 2.4.3 | |

**Supplementary materials, Table E16:** Productivity inputs

| Input | Value - males | Value - females | Source |
| --- | --- | --- | --- |
| Weekly wage 50 to 59 | £874.70 | £581.80 | ONS 2023: Table 6 (40)  Mean weekly wage |
| Weekly wage 60+ | £693.60 | £428.40 |  |
| Excess number of days absent due to smoking | 2 | | ONS 2020 (41) - Sheet 11b  Difference in annual number of sick days for current smokers (5.6) and those that have never smoked (3.6). |
| Proportion of smokers in employment | 45.7% | | ONS 2022 (17) |
| Average retirement age | 66 | | ONS 2024 (42) |

Abbreviations: ONS – Office for National Statistics.

Supplementary materials, Table E17: Summary of PSA distributions

| Parameter | PSA Distribution |
| --- | --- |
| Intervention effectiveness (RR) | NMA Coda* |
| Probability of abstinence (usual care – behavioural support only) | Beta [0,1] ^**^ |
| Smoking status (by age & gender)  Former smoker  Current smoker  Never-smoker | Beta [0,1] (Dirichlet)  Beta [0,1] (Dirichlet)  Beta [0,1] (Dirichlet) |
| Mortality per 1000 (by age & smoking status) | Beta [0,1000] |
| Comorbidities RR parameters  Stroke  Lung cancer  MI  CHD  COPD  Asthma | Log-normal  Log-normal  Log-normal  Log-normal  Log-normal  Log-normal |
| Comorbidities prevalence & incidence rates | Beta [0,1] |
| Utilities  Smoker/ former smoker/ non-smoker  CHD  All other comorbidities (excluding CHD) | Beta [0,1]  Beta [0,1]  Beta [0,1] |
| Intervention costs | Gamma |
| Comorbidity costs | Gamma |

Abbreviations: CHD – Coronary heart disease, COPD – Chronic obstructive pulmonary disorder, MI – Myocardial infarction, RR – Risk ratio.

*Results from the NMA Coda used to capture intervention effectiveness

**Values in square brackets indicate the limit of the beta distribution. For example, beta [0,1] indicates that a beta distribution is applied bounded between 0 and 1.

***Additional information on PSA***

PSA results (i.e. the probabilistic ICER and NMB) began to stabilise by around 1,000 iterations. Therefore, the PSA was run for 1,000 iterations, with weighted averages calculated within each iteration. Input parameter distributions for the PSA followed recommendations in Briggs et al. (2006) (29): beta distributions were applied to probabilities, prevalence rates and utilities; inverse normal distributions were applied to RR parameters; and gamma distributions were applied to costs. In addition, a (beta) Dirichlet distribution was applied to the age-related probabilities of being a current smoker, former smoker, and non-smoker to ensure the PSA values across these three parameters summed to one. The PSA distributions were fit using standard errors and 95% confidence intervals, or alpha (event rates) and beta (non-event rates) values, if these were available in the published literature i.e. reported alongside the mean estimates used to populate the base case model. If these were not available, then we applied an assumption that the value of the standard error was equal to 15% of the mean (base case) parameter value.

Supplementary materials, Table E18: Cost breakdown per person (lifetime)

| Intervention | Treatment costs | Stroke costs | Lung cancer costs | MI costs | CHD costs | COPD costs | Asthma costs | Private costs | Lost productivity costs |
| --- | --- | --- | --- | --- | --- | --- | --- | --- | --- |
| No intervention | £0.00 | £9,958 | £1,771 | £4,625 | £13,330 | £3,830 | £11.75 | £0.00 | £318.22 |
| Behavioural support only | £90.20 | £9,858 | £1,731 | £4,550 | £13,174 | £3,804 | £11.74 | £0.00 | £305.71 |
| NRT l/s | £167.58 | £9,690 | £1,663 | £4,423 | £13,080 | £3,760 | £11.72 | £0.00 | £284.59 |
| Cytisine | £205.20 | £9,644 | £1,644 | £4,388 | £13,054 | £3,747 | £11.72 | £130.91 | £278.73 |
| Varenicline | £272.14 | £9,601 | £1,627 | £4,356 | £13,030 | £3,736 | £11.71 | £0.00 | £273.40 |
| E-cigarettes | £213.76 | £9,605 | £1,629 | £4,359 | £13,033 | £3,737 | £11.71 | £0.00 | £273.90 |
| Varenicline + NRT l/s | £349.52 | £9,538 | £1,602 | £4,308 | £12,995 | £3,720 | £11.71 | £83.83 | £265.51 |
| NRT l&s | £211.04 | £9,512 | £1,591 | £4,288 | £12,981 | £3,713 | £11.70 | £83.83 | £262.20 |
| E-cigarettes + NRT l/s | £291.14 | £9,467 | £1,573 | £4,255 | £12,956 | £3,701 | £11.70 | £217.69 | £256.60 |

*	Figures are rounded, and therefore, there might be slight discrepancies in the totals.

**	Private costs and productivity costs were excluded from the ICER calculations presented in Table 3.1

Supplementary materials, Table E19: Incremental cost-effectiveness results (per person): fully incremental analysis (5-year time horizon)

| Intervention | RR vs. behavioural support only | 5-year costs | 5-year QALYs | ICER vs. behavioural support only | NMB vs. behavioural support only | CE rank |
| --- | --- | --- | --- | --- | --- | --- |
| No intervention | N/A | £8,302 | 3.22 | Dominated | -£100 | 9 |
| Behavioural support only | 1.00 | £8,277 | 3.22 | N/A | N/A | 8 |
| NRT l/s | 1.83 | £8,159 | 3.23 | Dominant | £243 | 7 |
| Cytisine | 2.06 | £8,142 | 3.23 | Dominant | £294 | 6 |
| Varenicline | 2.27 | £8,160 | 3.23 | Dominant | £308 | 5 |
| E-cigarettes | 2.25 | £8,106 | 3.23 | Dominant | £359 | 3 |
| Varenicline + NRT l/s | 2.58 | £8,164 | 3.23 | Dominant | £350 | 4 |
| NRT l&s | 2.71 | £7,995 | 3.23 | Dominant | £539 | 2 |
| E-cigarettes + NRT l/s | 2.93 | £8,024 | 3.23 | Dominant | £544 | 1 |

*	Figures are rounded, and therefore, there might be slight discrepancies in the totals.

Supplementary materials, Table E20: PSA results, cost-effectiveness threshold = £20,000

| Intervention | Probability cost-effective | |
| --- | --- | --- |
|  | Pairwise analysis  (Vs. behaviour support only) | Fully incremental analysis  (Vs. all other interventions) |
| Behavioural support only | N/A | 0% |
| NRT l/s | 100% | 0% |
| Cytisine | 100% | 1.8% |
| Varenicline | 100% | 0.3% |
| E-cigarettes | 99.4% | 8.5% |
| Varenicline + NRT l/s | 100% | 17.1% |
| NRT l&s | 100% | 24.7% |
| E-cigarettes + NRT l/s | 99.7% | 47.6% |

Supplementary materials, Table E21: Scenario assuming only difference in uptake for smoking cessation

| Result | Smoking cessation initiated at targeted lung screening programme | Usual care | Incremental |
| --- | --- | --- | --- |
| Costs per person | £32,779 | £33,276 | -£496 |
| QALYs per person | 9.09 | 9.02 | 0.07 |
| Private costs | £67 | £16 | £52 |
| Lost productivity costs | £284 | £310 | £26 |
| **ICER** | | | **Dominant** |
| **Net monetary benefit** | | | **£1,942** |

*	Figures are rounded, and therefore, there might be slight discrepancies in the totals.

Supplementary materials, Table E22: Scenario using three-year time horizon

| Result | Smoking cessation initiated at the lung cancer screening programme | Usual care | Incremental |
| --- | --- | --- | --- |
| Costs per person | £4,940 | £4,945 | -£5 |
| QALYs per person | 2.053 | 2.049 | 0.004 |
| Private costs | £67 | £1 | £67 |
| Lost productivity costs | £124 | £131 | -£13 |
| **ICER** | | | **Dominant** |
| **Net monetary benefit** | | | **£78** |

Abbreviations: ICER – Incremental cost-effectiveness ratio; QALY - Quality adjusted life year.

Supplementary materials, Table E23: Key modelling assumptions and rationale

| Assumption | Rationale |
| --- | --- |
| Where people in the cohort experienced multiple comorbidities, it was assumed that they experienced lowest utility of all of the morbidities which they experienced. | It is likely people with multiple co-morbidities would not have double the impact on their health, and there would be some interaction between conditions. Therefore, to apply more than one disutility is likely to overestimate the impact. However, we acknowledge applying only the most severe is a more conservative estimate, as it is likely to underestimate the impact. |
| Mortality is applied separately in the model, based on smoking status, and is not directly linked to the long-term conditions reflected in the model. | This is because smoking is likely to increase the risk of dying due to a range of different factors. By only capturing mortality associated with increased prevalence of long-term conditions will underestimate the impact of smoking on mortality. |
| Long-term conditions are captured using prevalence rates, and relative risks associated with smoking. Annual costs are then applied based on the prevalence rates. | This is a simplifying assumption as we are not modelling out the full pathway of long-term conditions. There is likely substantial heterogeneity in patient management of these conditions. Therefore, we are taking average costs associated with the condition. Furthermore, we also do not know people who survive or die due to the condition but have robust estimates of the prevalence by different age groups. |

Supplementary materials, Appendix A: PSA pairwise cost-effectiveness planes for all individual smoking cessation interventions

Cost-effectiveness plane: NRT I/s compared with behavioural support only

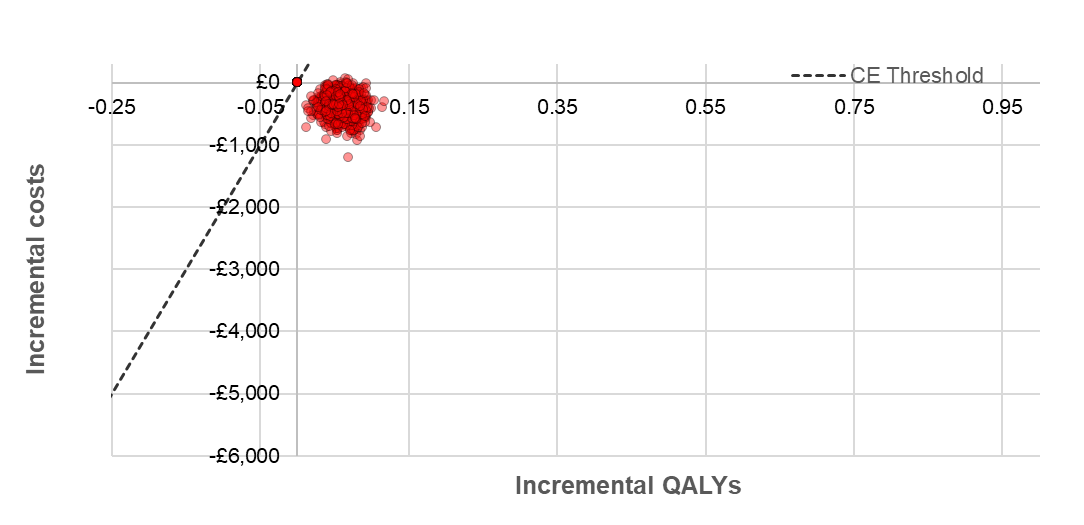

Cost-effectiveness plane: NRT I&s compared with behavioural support only

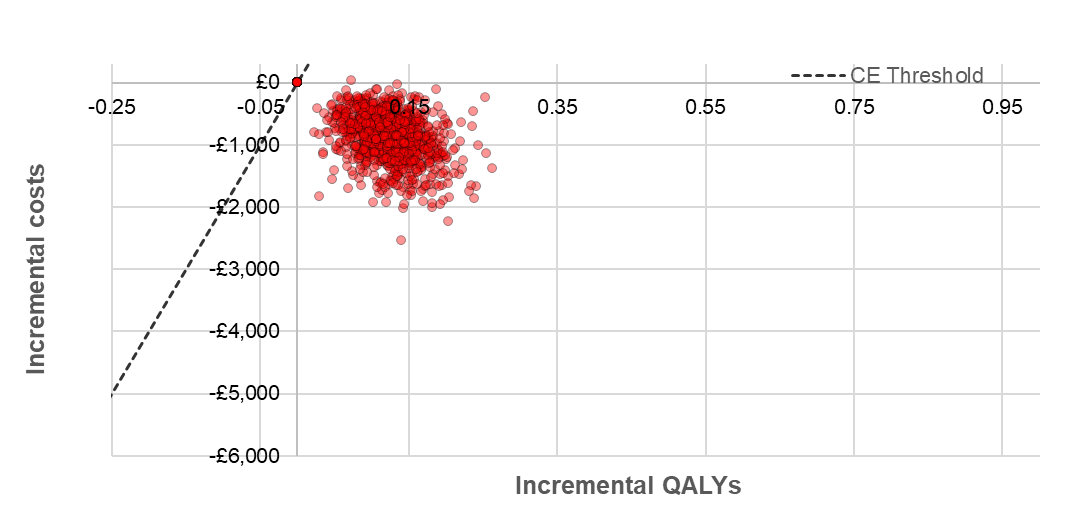

Cost-effectiveness plane: Cytisine compared with behavioural support only

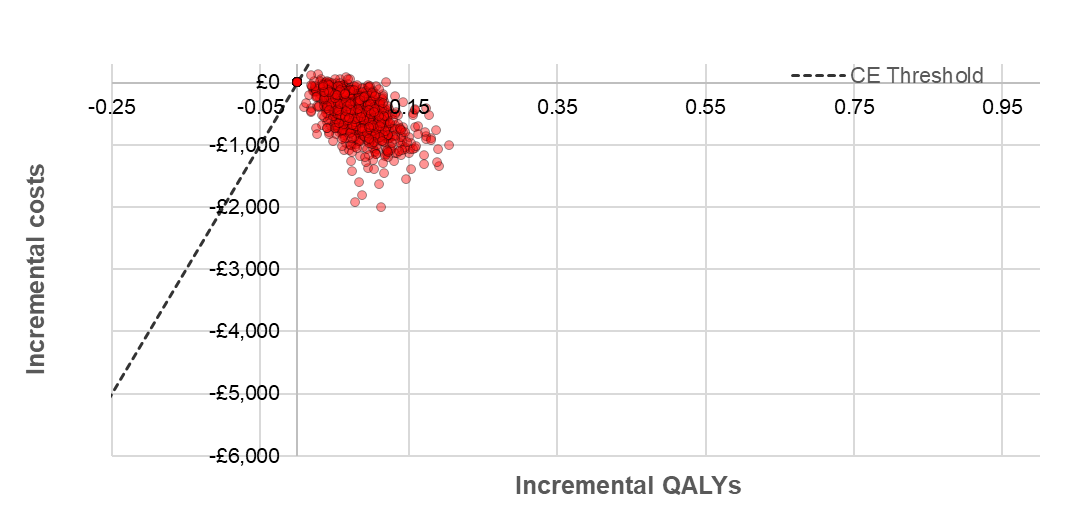

Cost-effectiveness plane: Varenicline compared with behavioural support only

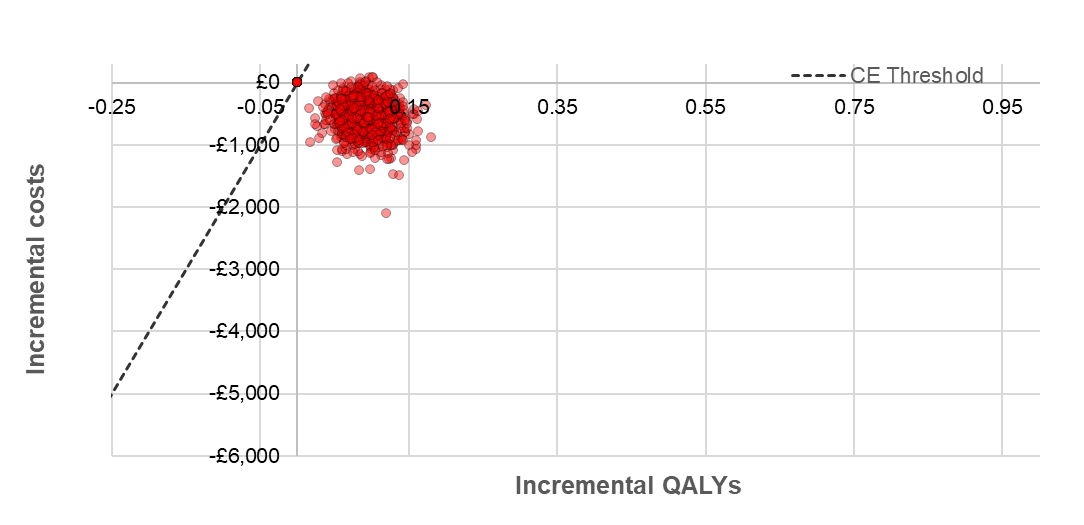

Cost-effectiveness plane: E-cigarettes compared with behavioural support only

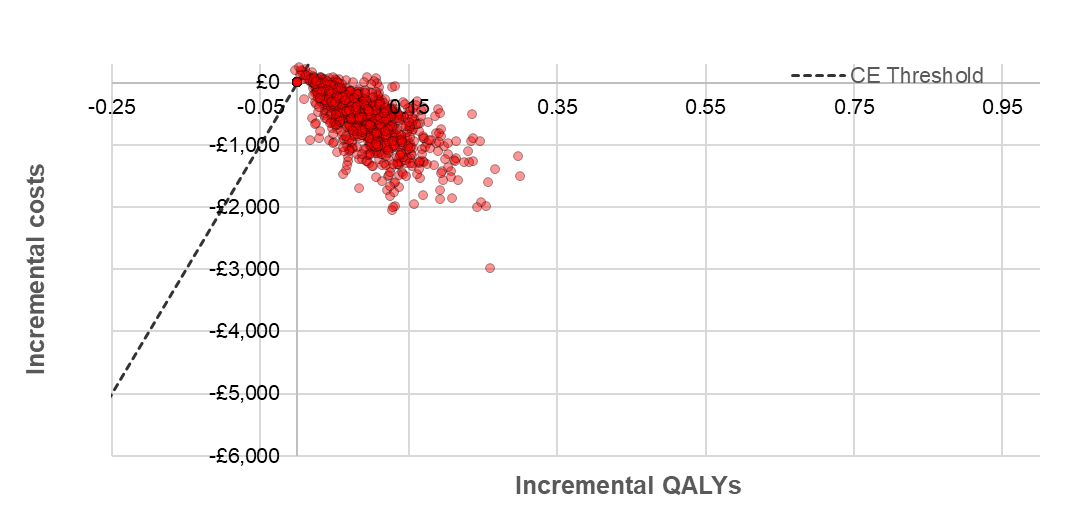

Cost-effectiveness plane: Varenicline + NRT I/s compared with behavioural support only

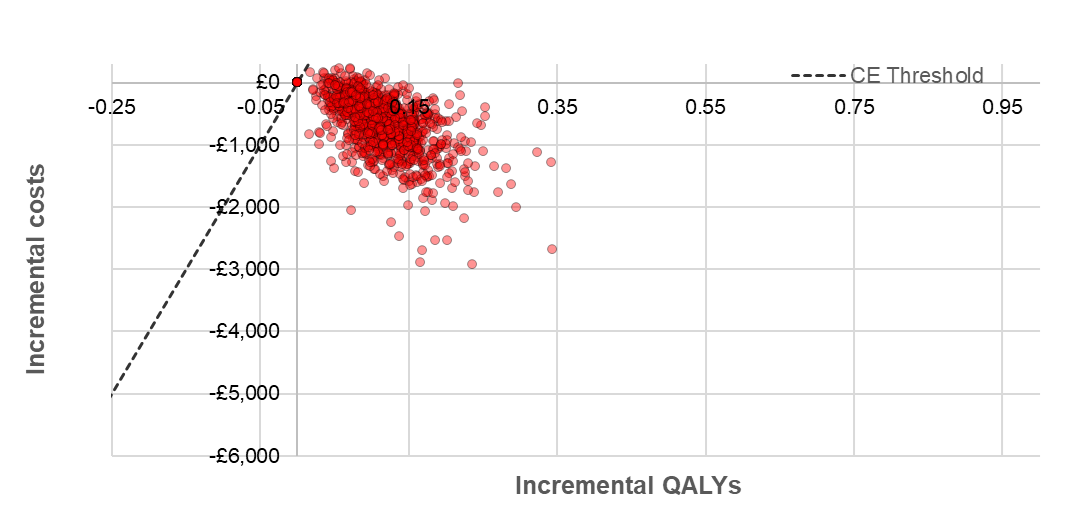

Cost-effectiveness plane: E-cigarettes + NRT I/s compared with behavioural support only

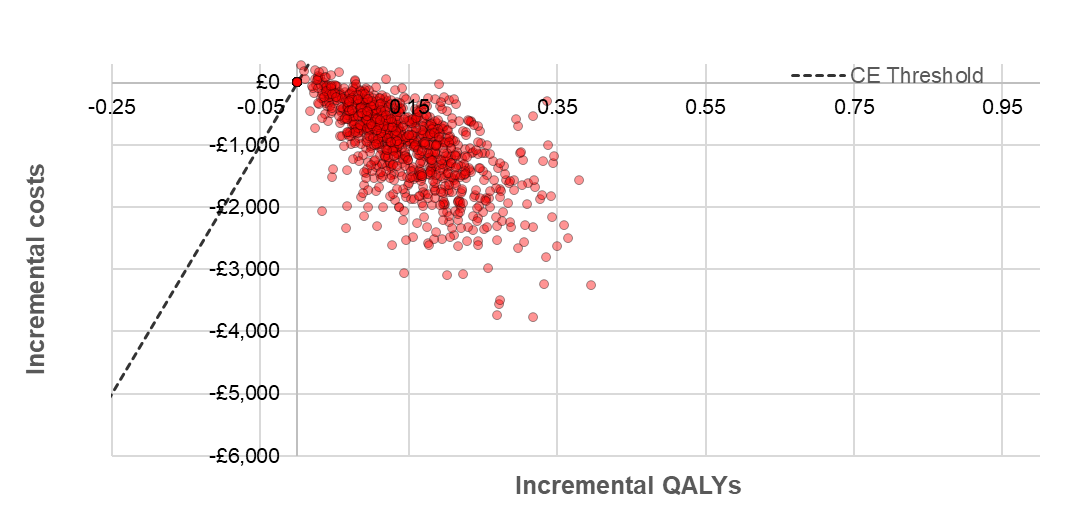

**Supplementary Materials References**

1.	Murray RL, Alexandris P, Baldwin D, Brain K, Britton J, Crosbie PAJ, et al. Uptake and 4-week quit rates from an opt-out co-located smoking cessation service delivered alongside community-based low-dose computed tomography screening within the Yorkshire Lung Screening Trial. European Respiratory Journal. 2024;63(4):2301768.

2.	Wu Q, Gilbert H, Nazareth I, Sutton S, Morris R, Petersen I, et al. Cost-effectiveness of personal tailored risk information and taster sessions to increase the uptake of the NHS stop smoking services: the Start2quit randomized controlled trial. Addiction. 2018;113(4):708-18.

3.	Williams PJ, Philip KEJ, Gill NK, Flannery D, Buttery S, Bartlett EC, et al. Immediate, Remote Smoking Cessation Intervention in Participants Undergoing a Targeted Lung Health Check: Quit Smoking Lung Health Intervention Trial, a Randomized Controlled Trial. Chest. 2023;163(2):455-63.

4.	National Institute for Health and Care Excellence. NG209: Economic Modelling Report 2018.

13.	National Health Service. NHS Prescription Cost Analysis 2023/24. 2024.

14.	Schnoll RA, Martinez E, Tatum KL, Glass M, Bernath A, Ferris D, et al. Increased self-efficacy to quit and perceived control over withdrawal symptoms predict smoking cessation following nicotine dependence treatment. Addict Behav. 2011;36(1-2):144-7.

15.	Personal Social Services Research Unit. The Unit Costs of Health and Social Care 2023 2024.

27.	NHS Digital. Health Survey for England, 2022. 2024.

28.	Bhatnagar P, Wickramasinghe K, Williams J, Rayner M, Townsend N. The epidemiology of cardiovascular disease in the UK 2014. Heart. 2015;101(15):1182-9.

29.	Myint PK, Sinha S, Luben RN, Bingham SA, Wareham NJ, Khaw KT. Risk factors for first-ever stroke in the EPIC-Norfolk prospective population-based study. Eur J Cardiovasc Prev Rehabil. 2008;15(6):663-9.

30.	Maddams J, Brewster D, Gavin A, Steward J, Elliott J, Utley M, et al. Cancer prevalence in the United Kingdom: estimates for 2008. British journal of cancer. 2009;101(3):541-7.

31.	O’Keeffe LM, Taylor G, Huxley RR, Mitchell P, Woodward M, Peters SAE. Smoking as a risk factor for lung cancer in women and men: a systematic review and meta-analysis. BMJ Open. 2018;8(10):e021611.

32.	NHS Digital. Health Survey for England, 2017. 2018.

33.	Millett ERC, Peters SAE, Woodward M. Sex differences in risk factors for myocardial infarction: cohort study of UK Biobank participants. BMJ. 2018;363:k4247.

34.	British Heart Foundation. Heart & Circulatory Disease Statistics 2024. 2024.

35.	Shields M, Wilkins K. Smoking, smoking cessation and heart disease risk: A 16-year follow-up study. Health reports. 2013;24(2):12-22.

36.	Public Health England. Estimated people with COPD. 2011.

37.	Forey BA, Thornton AJ, Lee PN. Systematic review with meta-analysis of the epidemiological evidence relating smoking to COPD, chronic bronchitis and emphysema. BMC Pulmonary Medicine. 2011;11(1):36.

38.	Doll R, Peto R, Wheatley K, Gray R, Sutherland I. Mortality in relation to smoking: 40 years' observations on male British doctors. BMJ. 1994;309(6959):901-11.

39.	Royal College of Physicians. Hiding in plain sight: Treating tobacco dependency in the NHS. 2018.

40.	Office for National Statistics. Earnings and hours worked, age group: ASHE Table 6. 2023.

41.	Office for National Statistics. Sickness absence in the UK labour market. 2023.

42.	Office for National Statistics. Milestones: journeying through modern life 2024 [Available from: <https://cy.ons.gov.uk/peoplepopulationandcommunity/populationandmigration/populationestimates/articles/milestonesjourneyingthroughmodernlife/2024-04-08#:~:text=Age%2066%3A%20Retiring&text=There%20has%20been%20a%20bigger,2011%20to%2066%20in%202021>.
